# Heterogeneous Genotype-Phenotype Associations in TRIO-Related Neurodevelopmental Disorder Revealed by Meta-Analysis

**DOI:** 10.1101/2025.10.17.25338222

**Authors:** Sarah Ann Duck, Alfred L. George

## Abstract

**Background:** The trio Rho guanine nucleotide exchange factor gene *(TRIO)* is highly expressed in the developing brain and contributes to neuronal development, specifically axon guidance, synaptogenesis, and cytoskeleton organization. Pathogenic *TRIO* variants are associated with a neurodevelopmental disorder with substantial phenotypic heterogeneity. Prior case series suggested genotype-phenotype associations related to variant type and location in the protein. However, these results need validation in a larger sample. The objectives of this research were to examine associations between phenotype, variant location and variant type among previously reported *TRIO*-related neurodevelopmental disorder cases, and identify recurrent *TRIO* variants.

**Methods:** Eighty-seven previously published studies annotated in the Human Gene Mutation Database reporting at least one *TRIO* variant were identified. Thirty-two additional cases were ascertained from the Simons Searchlight Study. A total of 699 individual case records were reviewed. After removing redundant cases, 449 unique records remained with available genotype data, of which 228 also had available phenotype information. Along with descriptive statistics, Chi-square analysis was used to test associations between variant and head size.

**Results:** In a meta-analysis of reported *TRIO* variants, categorically-defined head size is associated with variant type (missense vs truncating) and protein domain location (χ^2^ = 39.20; p = <0.001). Specifically, missense variants in the spectrin repeat domain are associated with macrocephaly whereas missense variants outside the spectrin domain and truncating variants are associated with microcephaly. The most prevalent phenotypic features were intellectual disability/developmental delay followed by autism spectrum disorder (ASD) or ASD-like behaviors. Seven recurrent *TRIO* variants were identified, with head size consistent across cases with the same variant.

**Conclusions:** *TRIO* variant type and location exhibit unique phenotypic associations. This observation may help clinicians and families to anticipate neurodevelopmental outcomes. Furthermore, identified recurrent variants may serve as targets for future translational and pharmacological research.

## Introduction

Neurodevelopmental disorders (NDD) encompass a heterogenous group of conditions associated with abnormal brain development impacting communication, behavior, motor skills, and cognition (1, 2). Approximately 16-18% of children are diagnosed with NDD, although reports range from 10-24% based on demographic factors (3-5). Autism spectrum disorder [ASD], intellectual disability [ID], and attention-deficit/hyperactivity disorder [ADHD] are prevalent examples of NDD (1, 2). NDD often present with comorbid conditions such as epilepsy (5-7) and may share genetic causes (6). Numerous genes have been implicated in the etiologies of NDD (8-11) including the *TRIO* gene (12). Identifying the genetic etiology of NDD can aid clinicians in the identification, treatment, and familial counseling of NDD (13).

The trio Rho guanine nucleotide exchange factor, encoded by *TRIO*, is highly expressed in the developing brain with major contributions to the development and function of neurons (12, 14, 15). Pathogenic *TRIO* variants are associated with a clinical phenotype broadly classified as *TRIO*-related NDD (16). Several small case series have documented the heterogenous phenotype of this disorder with common features including developmental delay, ID/ASD, ADHD, obsessive compulsive disorder, schizophrenia, seizures, facial dysmorphisms, and macrocephaly or microcephaly (12, 17-21). Beyond case reports, *TRIO* variants have also been identified in large population genetic analyses focused on NDD, although recurrent *TRIO* variants of have not been well delineated.

Case series have described a *TRIO* genotype-phenotype relationship. Specifically, a review paper summarizing 57 patient cases, reported that missense variants in the spectrin-repeat domain were associated with macrocephaly and severe ID, while missense variants in the GEFD1 and GEFD2 domains, truncating variants, and deletions were associated with microcephaly and less severe ID (18). However, given the small sample size, more research is needed to validate these findings in a larger sample.

We examined associations between phenotype, variant location and variant type in a meta-analysis of all previously reported *TRIO*-related NDD cases. This analysis also enables identification of recurrent *TRIO* pathogenic variants. Defining genotype-phenotype relationships unique to *TRIO* will help clinicians and families anticipate neurodevelopmental outcomes. Identification of recurrent variants delineates specific genetic etiologies of the disease that can be modeled in animals and cellular systems to investigate pathophysiology and conduct preclinical therapeutic development.

## Results

### Meta-Analysis

Eighty-seven previously published studies annotated in the Human Gene Mutation Database (Professional version 2025.2) as of June 2, 2025 were reviewed. A total of 667 individual cases were from the literature along with 32 more from the Simons Searchlight study, creating an initial cohort of 699 individual records for review. Forty records were excluded for missing or unclear genotype, 185 records were excluded for suspected duplicate individuals, and 25 records were excluded as data were not from an individual person, thus 449 unique records with genotype data were included in our analysis. Phenotype data that fell into any of six phenotypic categories was available for 228 out of the 449 cases. **Figure 1** presents a visual depiction of inclusion and exclusion of records.

**Figure 1.**
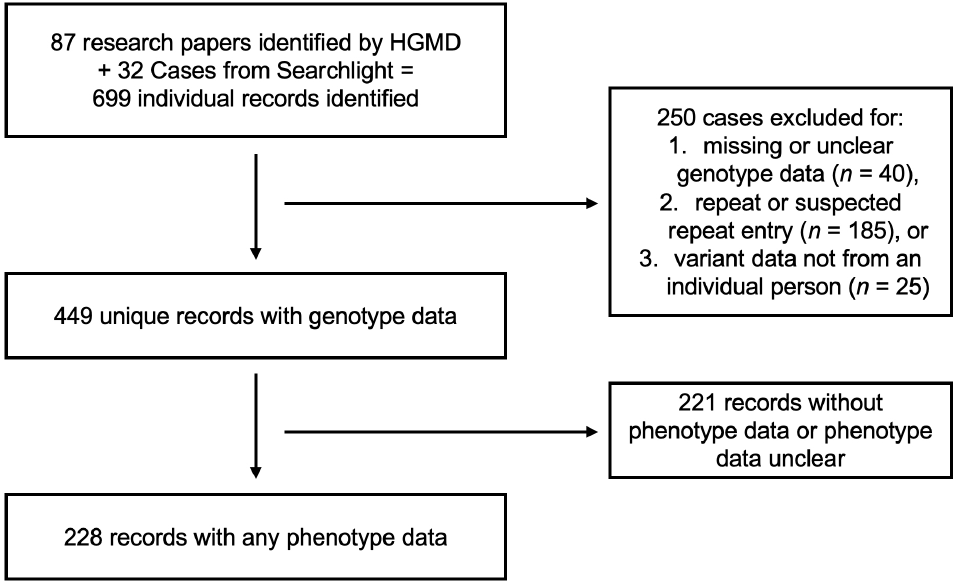
Flow diagram for selection of TRIO variant cases.

### Characteristics of Study Population

In the sample of 228 records with available phenotype data, 75 (32.9%) of individuals were female, 97 (42.5%) were male, and 56 (24.6%) had unknown sex. Variant classification was 152 (66.7%) missense, 57 (25%) truncating, and 19 (8.3%) other. Of participants with available head circumference data, 54 (53.4%) participants were classified as microcephalic, 22 (21.8%) macrocephalic, and 25 (24.8%) normocephalic. The most prevalent phenotypic features were ID/DD (*n* = 145) followed by ASD or ASD-like traits (*n*= 99). **Table 1** presents a summary of the study population.

**Table 1.** Study population characteristics.

|  |  | ***n* (%)** |
| --- | --- | --- |
| **Sex** | female | 75 (32.9%) |
|  | male | 97 (42.5%) |
|  | unknown | 56 (24.6%) |
| **Inheritance** | *de novo* | 67 (29.4%) |
|  | inherited | 33 (14.5%) |
|  | unknown | 128 (56.1%) |
| **Variant Type** | **Missense** |  |
|  | missense | 147 (64.5%) |
|  | in-frame insertion | 1 (0.4%) |
|  | in-frame deletion | 4 (1.8%) |
|  | **Truncating** |  |
|  | nonsense | 26 (11.4%) |
|  | frameshift | 29 (12.7%) |
|  | large deletions | 2 (0.8%) |
|  | **Other** |  |
|  | splice site | 10 (4.4%) |
|  | intron | 5 (2.2%) |
|  | synonymous | 4 (1.8%) |
| **Head Size** | microcephaly | 54 |
|  | macrocephaly | 22 |
|  | normocephaly | 25 |
| **Other** | seizure | 27 |
|  | ID/DD | 145 |
|  | ASD/ASD-like traits | 99 |
|  | ADHD/poor attention | 42 |
|  | schizophrenia | 7 |

*n* (%) = count (percentage); ID/DD = Intellectual Disability or Developmental Delay;
ASD = Autism Spectrum Disorder; ADHD = Attention-Deficit/Hyperactivity Disorder.

### In Genotype Phenotype Relationships

Categorically-defined head size was associated with variant location, and classification (χ^2^ = 39.20, p = <.001) with 16 (64%) individuals having a missense variant in the spectrin repeat domain exhibiting macrocephaly, and 23 (59%) individuals with a missense variant outside the spectrin repeat domain as well as 24 (75%) of those with a truncating variant exhibiting microcephaly (**Table 2a**). When examining variant classification in relationship to head size, missense variants are distributed evenly across the three phenotypes and truncating variants are primarily associated with microcephaly (χ^2^ = 12.20, p = 0.002; **Table 2b**). Variant location alone had a significant relationship with head size (χ^2^ = 26.43, p = <.001; **Table 2c**) with approximately half of the variants in the spectrin repeat domain associated with the macrocephaly and two-thirds of the variants in other regions of the protein associated with microcephaly. Variant phenotype mapped to the protein domains is depicted in **Figure 2**.

**Table 2.** Associations between head size with variant classification and domain location.

|  |  |  |  |  |
| --- | --- | --- | --- | --- |
| **2a.** |  |  |  |  |
| *Domain, Classification* | *Microcephaly* | *Macrocephaly* | *Normocephaly* | $\chi^2 = 39.20$, |
| Spectrin repeat, Missense | 2 (8.0%) | 16 (64.0%) | 7 (28.0%) | $p = <0.001$ |
| Not-spectrin, Missense | 23 (59.0%) | 4 (10.2%) | 12 (30.8%) |  |
| Truncating | 24 (75.0%) | 2 (6.2%) | 6 (18.8%) |  |
| **2b.** |  |  |  |  |
| *Classification* | *Microcephaly* | *Macrocephaly* | $\chi^2 = 12.20$, | $\chi^2 = 12.20$, |
| Missense | 25 (39.1%) | 20 (31.3%) | 19 (29.6%) | $p = 0.002$ |
| Truncating | 24 (75.0%) | 2 (6.2%) | 6 (18.8%) |  |
| **2c.** |  |  |  |  |
| *Domain* | *Microcephaly* | *Macrocephaly* | *Normocephaly* | $\chi^2 = 26.43$, |
| Spectrin repeat | 9 (27.3%) | 17 (51.5%) | 7 (21.2%) | $p = <0.001$ |
| Non-Spectrin | 45 (66.2%) | 5 (7.3%) | 18 (26.5%) |  |

data displayed as count (percentage); $\chi^2$ = chi-square.

**Figure 2.**
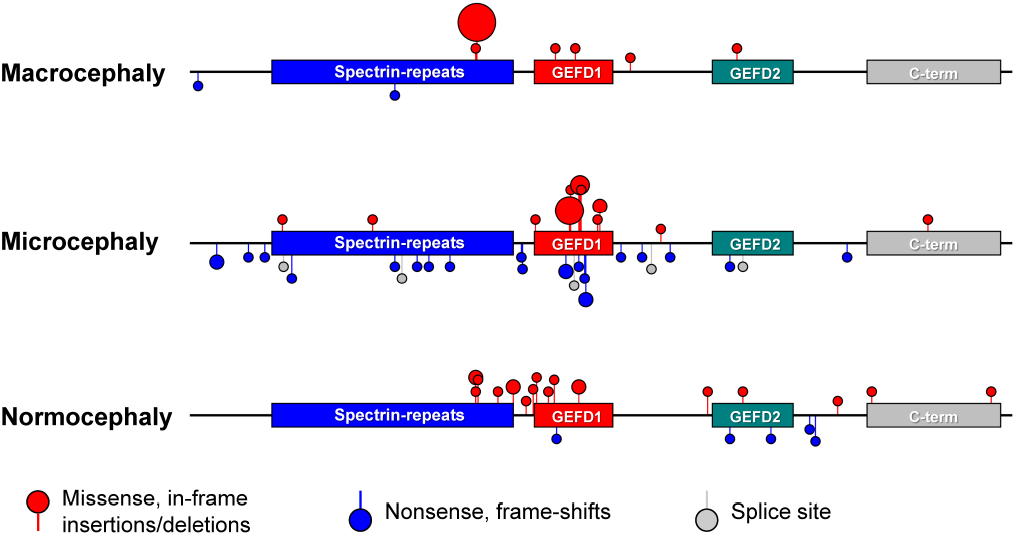
TRIO Variant Location grouped by head size phenotype. Individual lollipops mark positions of variant codons with missense (red) variants projecting above, and truncating (blue) and splice site (grey) variants projecting below the linear depiction of the TRIO protein. The symbol size is approximately proportional to the number of variants at each position.

Additionally, we examined genotype associations with neurological phenotypes, including seizures and ID/DD (**Figure 3**). Variants associated with seizures were distributed across protein domains with the majority (13; 81.3%) of missense variants occurring in the spectrin repeat domain and all truncating variants (10; 100%) occurring outside of this domain. Variants associated with ID/DD were both missense and truncating distributed throughout the protein. All (15; 100%) truncating variants associated with ASD and ASD-like traits occurred outside of the spectrin repeat domain, while missense variants were distributed throughout the protein. Furthermore, 73 (83%) variants associated with ASD/ASD-like traits were classified as missense, while only 15 (17%) were truncating. Missense (19; 51.4%) and truncating (18; 48.6%) variants were equally associated with ADHD and poor attention and were distributed throughout the protein. Schizophrenia was the least prevalent phenotype observed in our sample. One missense variant was associated with schizophrenia and occurred at the end of the spectrin repeat domain. Four truncating variants associated with schizophrenia were distributed throughout the rest of the protein.

**Figure 3.**
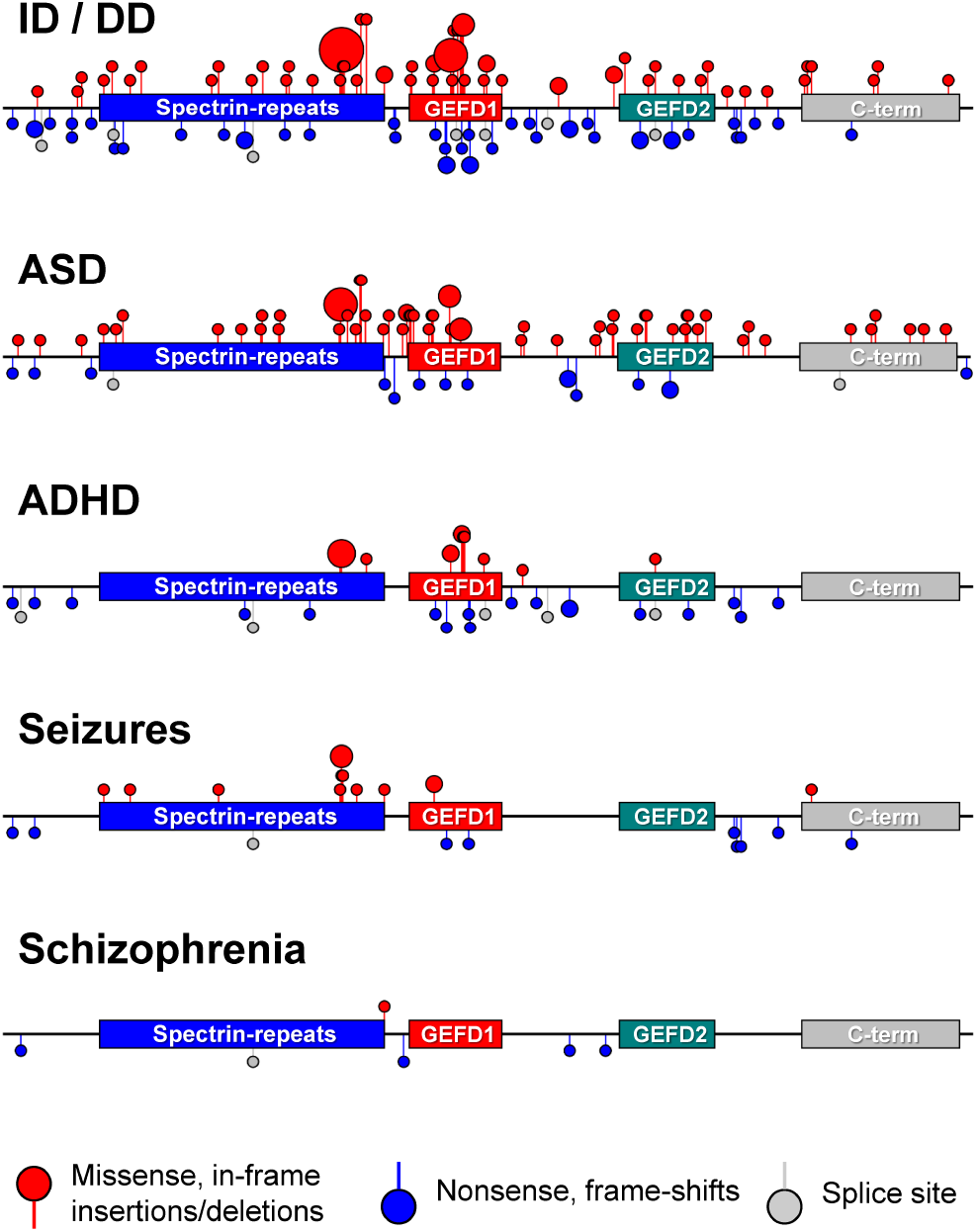
TRIO variant location grouped by phenotype. Individual lollipops mark positions of variant codons with missense (red) variants projecting above, and truncating (blue) and splice site (grey) variants projecting below the linear depiction of the TRIO protein. The symbol size is approximately proportional to the number of variants at each position.

### Recurrent *TRIO* Variants

We identified 7 missense variants with 5 or more recurrences (Arg1078Trp, Arg1078Gln, Arg1428Gln, Pro1461Leu, Asn1465Ser, Ser2767Leu, and Gly3053Ser). We observed no recurrent truncating variants. Four of the recurrent missense variants occur in context of CpG dinucleotides (Arg1078Trp, Arg1078Gln, Arg1428Gln, Pro1461Leu). Head size was consistent among individuals with a given recurrent variant. Specifically, Arg1078Trp and Arg1078Gln were associated with macrocephaly, Arg1428Gln and Asn1465Ser were associated with microcephaly, and Pro1462Leu was associated with normocephaly. Recurrent variants Gly3053Ser and Ser2767Leu did not have available phenotype information.

## Discussion

*TRIO* is highly conserved among different species (26), with complete knockout in mice being lethal (15, 27). The TRIO protein product consists of multiple distinct domains including spectrin-like repeats, two guanine nucleotide exchange factor domains (GEFD1 and GEFD2), and a C-terminal kinase domain. GEFD1 activates the small GTPases Rac1 and RhoG, while GEFD2 activates RhoA (12, 28, 29). TRIO is implicated in synaptic structure and function (12, 30-33), axon guidance (34-37) and cell adhesion (26, 38).

In this study, we investigated genotype-phenotype relationships in *TRIO*-related NDD using a meta-analysis of previously reported-cases including a survey of recurrent variants in this gene. In our sample of 228 unique records with available phenotype data, we found ID/DD to be the most prevalent phenotype followed by ASD/ASD-like traits, ADHD/poor attention, seizures, with only a few cases of schizophrenia. The predominance of neurological and neuropsychiatric phenotypes in our sample is mechanistically consistent with previous research that has established the role of *TRIO* in brain development and neuron function (12, 14, 15, 28-31).

Previous case series noted associations of macrocephaly and microcephaly with pathogenic variants in specific protein domains (16, 18). We also observed these associations in our larger study population, but they were less absolute. Specifically, individuals with missense variants in the spectrin repeat domain were statistically more likely to exhibit macrocephaly, while those with truncating variants or missense variants outside of the spectrin repeat domain were more likely to exhibit microcephaly. Genetic background or interaction with the environment through epigenetic modification may influence the ultimate phenotype. However, head size was consistent within recurrent variants, for those with phenotype information available, which may indicate that differences in a domain are due to gain or loss of function conferred by each unique variant, which would warrant further testing.

As clinical genetic testing becomes more routine, understanding trends in the genotype-phenotype relationship of *TRIO*-related NDD may help families anticipate neurodevelopmental outcomes and provide physicians with an expected clinical course and prognosis. Identifying a specific genetic variant can promote more individualized therapeutic and behavioral care plans.

While many genotypes observed in our sample were unique, there were several recurrent variants. Four of the 7 identified missense recurrent variants were in the context of CpG dinucleotides which is a common hotspot for mutation events (39, 40). Novel gene therapy and other pharmaceutical research may benefit from knowing these recurrent variants for establishing animal and cellular models to test candidate treatments as they may represent a larger proportion of the *TRIO*-related NDD population and span several *TRIO* domains.

### Study Limitations

The main limitation of this study is incomplete phenotypic information and the retrospective nature of the data collection. Many reports did not report on all six of the phenotypic categories that we investigated, and we purposely avoided over interpretation of absent information as evidence of absent phenotypes. Therefore, there was no negative control group available within the sample for any of the phenotypic categories, other than quantitative head circumference. Although individuals clearly reported in multiple publications were counted only once in our study, there may remain redundant entries despite our best efforts to remove all confirmed or suspected duplicate individuals. Thus, we set the threshold for defining a recurrent variant at 5 or more recurrences to account for that possibility.

## Conclusion

In conclusion, *TRIO* variant type and location exhibit unique phenotypic associations. This observation may help clinicians and families to anticipate neurodevelopmental outcomes. Furthermore, identified recurrent variants may serve as targets for future translational and pharmacological research.

## Data Availability

Data from the Simons Searchlight study are available by direct request to the Simons Foundation (https://base.sfari.org). All other data generated from meta-analysis are included in this article and its supplementary information files.

## Ethics Approval and Consent to Participate

The Northwestern University Institutional Review Board classified access to deidentified data in Simons Searchlight as not human subjects research.

## Authors’ Contributions

S.D. and A.G. contributed to the conceptualization, methodology, visualization, manuscript writing, and review & editing. A.G. supervised the project and S.D. conducted formal analysis and investigation.

## Acknowledgements

The authors are grateful for access to the SFARI data and to all of the families who participated in the Simons Searchlight Project. We thank Dr. Katina Calakos for help navigating the Simons Searchlight data. We also thank Jean-Marc DeKeyser for developing the lollipop plotting software.

## Methods

### Meta Analysis

Publications that reported at least 1 *TRIO* variant were identified using the Human Gene Mutation Database (Professional version 2025.2) (22). Individual variant records were extracted from the literature as well as cases ascertained from the Simons Searchlight Study (dataset version 13). To identify unique variant records with genotype data, the following exclusion criteria were applied: 1) missing or unclear genotype data, 2) suspected duplicate individual, 3) variant data not from an individual person. Duplicate individuals were determined using direct references to original publications included in case presentations, common patient identifiers from larger publicly accessible databases, and overlapping data sources as detailed in published methods sections. When unclear, records were counted as a duplicate to limit redundancy as much as possible. Phenotype data that fell into any of six a priori phenotypic categories (head size, seizures, intellectual disability or developmental delay [ID/DD], autism spectrum disorder [ASD] or ASD-like traits, attention-deficit/hyperactivity disorder [ADHD] or poor attention, and schizophrenia) was recorded, as well as patient sex, variant inheritance, and variant type. Head size was categorized into macrocephaly, microcephaly and normocephaly based on reported head circumference.

Thirty-two participants in the Simons Searchlight study were ascertained from the Simons Foundation Autism Research Initiative (SFARI) database (23). Simons Searchlight is a registry that recruits participants with known genotype in approximately 180 genes associated with neurodevelopmental disorders. The study collected longitudinal natural history data including medical history, seizure history, and psychometric testing. Simons Searchlight reviewed individual genetic test reports as previously described (23, 24). For this study, 31 participants with *TRIO* variants were included as well as 1 proband (SFARI ID: 17025-x2) with a secondary finding of a *TRIO* pathogenic variant. For all participants, variant genotype information was obtained using data from “searchlight_core_descriptive_variables”, version 13 (released July 2025). Eighteen participants identified as probands had available phenotype information. Head size and seizure information was extracted from “svip_mhi_summary” while ASD, ID/DD, ADHD, and schizophrenia spectrum was extracted from “previous_diagnoses_summary.” The Northwestern University Institutional Review Board classified access to deidentified data in Simons Searchlight as not human subjects research.

### Data Analysis

Variant classification was determined and parsed into three groups: 1) missense variants, consisting of missense variants, small in-frame insertions, and small in-frame deletions, 2) truncating variants, consisting of nonsense variants, frameshift variants, and large deletions, and 3) other variants, consisting of splice site, intron, and synonymous variants. If only protein nomenclature or cDNA nomenclature was available, the missing genotype data (i.e. protein or cDNA nomenclature) was added based on the reference sequence NM_007118. Variant location in the *TRIO* protein was assigned using domain definitions from Uniprot (25). When occipitofrontal circumference was available as a measure of head size, macrocephaly was defined as greater than 2 standard deviations (SD) above the mean; microcephaly, less than 2 SD; and normocephaly, between negative and positive 2 SD. Along with descriptive statistics, Chi-square tests were used to analyze associations between variant and head size, with p < 0.05 deemed statistically significant. The remaining phenotype data were presented with descriptive statistics. Because most publications recorded only the presence not absence of a phenotype, there was no negative control group within the dataset. Recurrent *TRIO* variants were identified using the complete sample of non-duplicative records with available genotype data (N = 449). For determining the number of recurrent variants, only one person from the same family were counted. We used a frequency of greater than or equal to 5 recurrences as a cutoff point to account for any unidentified repeat cases within the dataset.

